# Recovery Time and Its Predictors among Women Admitted with Obstetric Fistula in Jimma University Medical Center Southwest, Ethiopia: A Retrospective Cohort Study

**DOI:** 10.1101/2022.02.02.22270299

**Authors:** Abriham Shiferaw Areba, Geremew Muleta Akessa, Megersa Tadesse, Arega Haile, Belayneh Genoro Abire, Aneleay Cherinet Eritero, Belay Negassa Gondol, Denebo Ersulo Akiso, Girum Gebremeskel Kanno, Desta Erkalo Abame, Negasa Eshete Soboksa

## Abstract

**Background:** Obstetric fistula is one of the most shocking medical disabilities that women face as a result of problems caused by a lack of surgical intervention.

**Methods:** From January 2011 to February 2017, a retrospective analysis was undertaken on obstetric fistula participants who received undergone surgical repair at the Jimma University Medical Center.

**Results:** 81.4% of fistula patients were reported to have recovered. The results of lognormal inverse-Gaussian frailty model revealed that weight ≥50kg (Ф= 0.86, 95% CI: 0.747, 0.99), divorced (Ф=1.2, 95% CI: 1.028, 1.413), urban residence (Ф= 1.56, 95% CI: 01.046, 2.317), urine incontinence >3 months (Ф=1.2, 95% CI: 1.056, 1.371), no antenatal care (Ф=1.189, 95% CI:1.023, 1.382), duration of labor <2days (Ф= 0.86, 95% CI: 0.758, 0.986), health center delivery (Ф= 0.76, 95% CI: 0.576, 0.989), vaginal delivery (Ф= 1.5, 95% CI: 1.125, 1.996), and partially damaged urethra (Ф=1.59, 95% CI: 1.168, 2.166) and recto-vaginal fistula(Ф= 0.735, 95%CI: 0.6045, 0.894) were associated with the recovery time.

**Conclusion:** Patients with a weight < 50 kg, delivery at a health center, labor lasting <2 days, urinary incontinence lasting <3 months, antenatal care follow-up, cesarean section delivery, urethra status not damaged, and recto-vaginal fistula had a faster recovery period.

## Background

An obstetric fistula is an improper connection between the vagina, rectum, and/or bladder that can form during a long and obstructed childbirth and cause persistent urine or fecal incontinence, according to the World Health Organization. Obstructed labor is one of the primary causes of maternal mortality in underdeveloped nations, and it comes with a slew of other complications, the most serious of which is obstetric fistula. For every maternal mortality, 20-30 women are projected to experience major obstetric problems, including fistula. Apart from surviving the experience of obstructed labor, these women must also deal with the physical and psychological obstacles of living with an obstetric fistula (1).

Obstetric fistula can occur in any developing country, although it is most common in the “fistula belt” that runs from Mauritania to Eritrea in the northern part of Sub-Saharan Africa, as well as the emerging countries of the Middle East and Asia (2). Though the exact prevalence is difficult to quantify, it is hypothesized that patterns follow maternal mortality ratios, with higher prevalence seen in areas where maternal mortality is high. In contrast, in affluent countries, obstetric fistula is mostly caused by iatrogenic factors such as radiation therapy and surgery (3).

In Ethiopia, where 74% of deliveries take place at home without medical assistance, the danger of death and fistulas for women giving birth is quite high. An estimated 8,500 to 9,000 of Ethiopia’s 3 million women who give birth each year may develop an obstetric fistula. Obstetrics accounts for 95 percent of VVF in Ethiopia. Obstructed labor that is not alleviated in time by conducting a cesarean section is the main cause of over 85 percent of OBF (4).

## Methods

### Study setting and design

A retrospective patients who had their fistula repaired at Jimma University Medical Center between January 2011 and February 2017 based on data from the OBF. From the date of follow-up entry, data was taken from the patient’s chart of all obstetric fistula patients. Only 270 women with OBF patients whose cards contained complete information were found from a total of 350 OBF patients enrolled in the given year, and data analysis was performed using statistical software R tools.

### Variable in the Study

Time to recovery from obstetric fistula, OBF diagnostic recovery, or censored (1^st^ January 2011 to 1^st^ February 2017) is the outcome variable, which is assessed in weeks. The age of occurrence, weight, marital status, parity, location of residency, education level, duration of urinary incontinence, ANC, labor, delivery place, delivery mode, surgery approach, catheter, urethral status, and forms of fistula are the explanatory variables in this study.

### Measurement of Variables

#### Recovered

are those patients who have become free from medical complications.

#### Recovery time

number of weeks it takes from date of entry until a patient is recovered from OBF. (The length of time between the date of entry and discharge).

#### Survival analysis

is a statistical method for data analysis where the outcome variable of interest is the time to the occurrence of an event. It is also referred to as “time-to-event analysis”

#### Survival time

describes the time from a certain origin to the occurrence of an event.

#### Censoring

occurs when we have some information about individual survival time, but we don’t know the survival time exactly.

### Data Analysis Method

For descriptive analysis, the data was evaluated using frequency and percentage. For inferential analysis, survival analysis is also known as “time-to-event analysis.” With censored data, things get a little more complicated (6). The Kaplan-Meier estimator is a non-parametric estimator that uses graphical methods to visualize data, and the log-rank test is a non-parametric test that compares two or more groups on the same Kaplan-Meier curves (7). The Cox PH regression model for the hazard function is a prominent semi-parametric survival model that allows the insertion of covariates while leaving the baseline risks undetermined and taking only positive values (9).

### Testing the Assumption of Cox PH Model

The assumptions are checked by variables in the final model. If the newly added variable is not significant, it can be taken as the assumptions of the proportional hazard assumptions are satisfied. The residuals constructed for each covariate that is included in the model which are expected to predict the recovery time of patient with obstetric fistula (10). If this plot shows some trend the assumption is violated, whereas if the plot demonstrates random distribution around the reference line then the assumption is satisfied.

The AFT model is a parametric survival model. The members of the AFT model considered in this study are the Weibull, log-logistic, and log-normal AFT models (11). Parametric Shared By using the zone of patients as the clustering variable, a frailty survival model was applied to uncover critical variables of recovery time after OBF. The frailty term’s value is constant throughout time and is shared by all members in the cluster, therefore it’s what causes the cluster’s event times to be dependent on one another (12, 13). For model comparison, Akaike’s information criterion was utilized. The statistical package R was used for data cleaning, management, and analysis (14).

## Results

The descriptive results of the variables for OBF during the therapy period are reported in this section. 81.4% of the OBF patients in the research were recovered, while the remaining 18.6% were censored.

### Comparison of Survival Experiences of OBF Patients

Figure 1 shows a survival curve of time-to-recovery based on incontinence duration and antenatal care. The data demonstrate that among patients with varying duration of incontinence and antenatal care, those with incontinence of less than three months and antenatal care follow-up had a faster recovery than those with incontinence of more than three months and no antenatal care service. This impression was verified by formal hypothesis tests, as shown in table 1; log-rank tests reveal a significant difference in recovery time (P = 3.16e-06, 0.0023).

**Figure 1:**
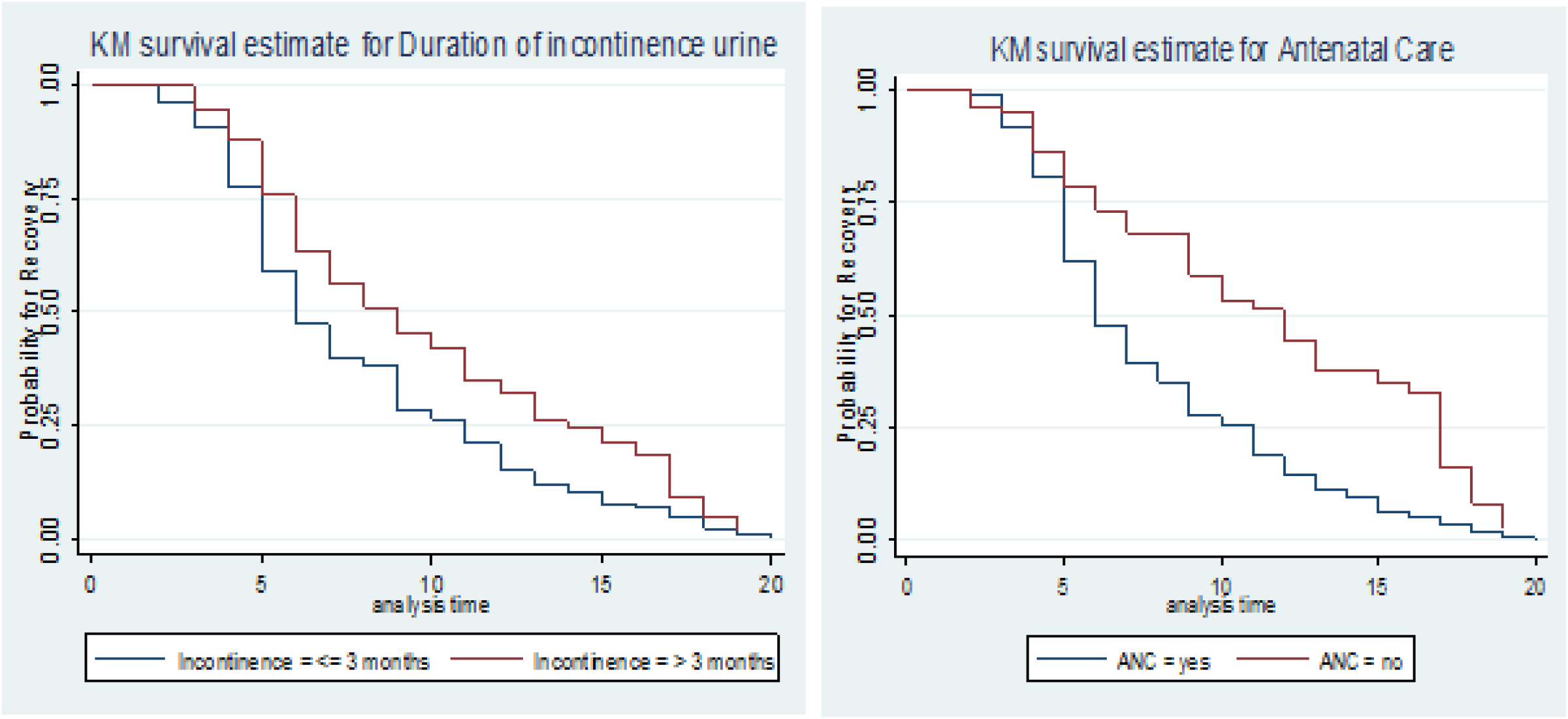
Kaplan Meier survivor estimates for categorical variables

**Table 1:** Summary results and log-rank test for survival time among covariates for OBF in JUMC

| Covariates | Categories | Frequency | Percent | Log Rank test |  |
| --- | --- | --- | --- | --- | --- |
| | | | | $\chi^2$ | P-value |
| Age | <20 | 67 | 24.8 | 0.1 | 0.957 |
|  | 21-30 | 137 | 50.8 |  |  |
|  | >30 | 66 | 24.4 |  |  |
| Weight | <50 kg | 197 | 73 | 18.3 | 1.84e-05 |
|  | ≥50 kg | 73 | 27 |  |  |
| Marital status | Married | 171 | 63.4 | 19.4 | 6.04e-05 |
|  | Divorced | 90 | 33.3 |  |  |
|  | Others | 9 | 3.3 |  |  |
| Parity | 1 Child | 83 | 30.7 | 7.3 | 0.0263 |
|  | 2-4 Children | 101 | 37.4 |  |  |
|  | >5 Children | 86 | 31.9 |  |  |
| Place of Residence | Rural | 259 | 95.9 | 16.7 | 4.29e-05 |
|  | Urban | 11 | 4.1 |  |  |
| Education Level | Illiterate | 179 | 66.7 | 0.3 | 0.607 |
|  | Literate | 91 | 33.3 |  |  |
| Duration of Incontinence of urine | >3 Months | 136 | 50.4 | 9.3 | 0.00226 |
|  | ≥ Months | 134 | 49.6 |  |  |
| Antenatal Care | No | 172 | 63.7 | 21.7 | 3.16e-06 |
|  | Yes | 98 | 36.3 |  |  |
| Duration of Labor | <2 Days | 83 | 30.7 | 2 | 0.157 |
|  | ≥2 Days | 187 | 69.3 |  |  |
| Place of Delivery | Home | 177 | 65.6 | 4.2 | 0.0395 |
|  | Health center | 93 | 34.4 |  |  |
| Mode of Delivery | Vaginal | 182 | 67.4 | 9.4 | 0.00217 |
|  | Cesarean section | 88 | 32.6 |  |  |
| Surgery Approach | Vaginal | 183 | 67.8 | 6.3 | 0.0121 |
|  | Abdominal | 87 | 32.2 |  |  |
| Duration of Catheter | ≤ 14 Days | 201 | 74.4 | 3.6 | 0.0593 |
|  | >21 Days | 69 | 25.6 |  |  |
| Status of Urethra | Not damaged | 214 | 79.3 | 20.1 | 7.52e-06 |
|  | Partially damaged | 56 | 20.7 |  |  |
| Types of fistula | RVF | 34 | 12.6 | 17.6 | 2.77e-05 |
|  | VVF | 236 | 87.4 |  |  |

**Table 2:** Results of multivariable Cox PH Model time-to-recovery from OBF in JUMC

| Covariates | Category | $\hat{\beta}$ | SE | Wald | P-value | HR | 95% CI for HR |
| --- | --- | --- | --- | --- | --- | --- | --- |
| Weight | < 50kg® |  |  |  |  |  |  |
|  | ≥50kg | 0.412 | 0.159 | 2.59 | 0.0095 | 1.51 | [1.11, 2.06] |
| Place of Residence | Rural® |  |  |  |  |  |  |
|  | Urban | -0.97 | 0.457 | -2.12 | 0.034 | 0.379 | [0.155, 0.93] |
| Incontinence of Urine | ≤3 months® |  |  |  |  |  |  |
|  | > 3 months | -0.297 | 0.147 | -2.02 | 0.044 | 0.743 | [0.556, 0.992] |
| Antenatal care | Yes® |  |  |  |  |  |  |
|  | No | -0.35 | 0.175 | -1.99 | 0.046 | 0.71 | [0.50, 0.994] |
| Mode of delivery | Cesarean section® |  |  |  |  |  |  |
|  | Vaginal | -0.576 | 0.28 | -2.06 | 0.04 | 0.562 | [0.325, 0.97] |
| Status of Urethra | not P. damaged® |  |  |  |  |  |  |
|  | P. damaged | -0.978 | 0.406 | -2.41 | 0.016 | 0.376 | [0.1696, 0.83] |

### Cox proportional Hazards Regression Model

A single covariate Cox proportional hazards model analysis is a good way to screen out potentially essential variables before putting them into the multivariable model. Weight of women, marital status, parity, place of residence, duration of urinary incontinence, antenatal care, duration of labor, place of delivery, mode of delivery, surgery approach, duration of catheter, status of urethra, and types of fistula were all significantly related to patient survival time, but age and level of education were not significant at a modest level of significance of 0.25.

### Multivariable Analysis of Cox PH Regression Model

The p-value associated with each parameter was assessed to determine whether or not a variable is significant, and variables with a p-value less than 0.05 were considered relevant variables and so included in the study.

## Assessment of Model Adequacy of Cox Proportional Hazards (PH) Model

The PH model’s assumptions are shown in Table 3. Except for the place of residence, all independent variables in the model violate the PH assumption for those covariates, as newly introduced covariates were statistically significant, indicating that proportional hazard assumptions were not met.

**Table 3:** PH assumption the predictor variables & their interaction with log time for time-to-recovery from OBF in JUMC

| Covariates | $\hat{\beta}$ | HR | SE | Wald | P-value |
| --- | --- | --- | --- | --- | --- |
| Weight | 3.4427 | 31.27 | 0.6988 | 4.93 | 8.4e-07 |
| Place of Residence | 1.966 | 7.144 | 2.9361 | 0.67 | 0.50308 |
| Incontinence of Urine | 2.884 | 17.8765 | 0.5996 | 4.81 | 0.50308 |
| Antenatal care | 3.336 | 28.102 | 0.6496 | 5.14 | 1.5e-06 |
| Mode of delivery | 2.437 | 11.441 | 0.6913 | 3.53 | 0.00042 |
| Status of Urethra | -5.844 | 0.0029 | 1.6532 | -3.53 | 0.00041 |
| Weight*ln(t) | -1.583 | 0.205 | 0.3514 | -4.51 | 6.6e-06 |
| Place of Residence *ln(t) | -1.182 | 0.3066 | 1.0812 | -1.09 | 0.27419 |
| Incontinence of Urine*ln(t) | -1.373 | 0.2535 | 0.2715 | -5.06 | 4.3e-07 |
| Antenatal care*ln(t) | -1.582 | 0.2056 | 0.2926 | -5.41 | 6.4e-08 |
| Mode of delivery*ln(t) | -1.196 | 0.3024 | 0.3182 | -3.76 | 0.00017 |
| Status of Urethra*ln(t) | 1.885 | 6.5837 | 0.6822 | 2.76 | 0.00574 |

### Accelerated Failure Time Model Results

Based on AIC, a model with the lowest AIC value was picked for the survival time of OBF patients’ data. From the options shown in table 4, the lognormal AFT model (AIC = 1206) was found to be the best for the survival time of the OBF data set.

**Table 4:** AIC value of parametric AFT model for time-to-recovery from OBF in JUMC

| Baseline Distribution | AIC |
| --- | --- |
| Weibull | 1223.792 |
| Lognormal | 1206.000* |
| Log- logistic | 1210.215 |

Under the lognormal AFT model showed above table 5, patients ’ weight ≥ 50kg is estimated to be 0.86 with [95% CI: 0.748, 0.992] by using <50kg as the reference category. The result shows that patient who had weight greater than/ equal to 50kg were short time-to-recovery from obstetric fistula than weight less than 50 kilogram. The acceleration factor for divorced patients was estimated to be 1.21. Divorce patients had a prolonged survival time of OBF than married patients. Acceleration factor of patients for the duration of labor less than 2days was 0.87 with [95% CI: 0.758, 0.987, and P=0.0389] by using labor ≥ 2days as the reference category. This implied that those patient who had the duration of labor less than 2days is fast time-to-recovery from OBF than the duration of labor ≥ 2days.

**Table 5:**
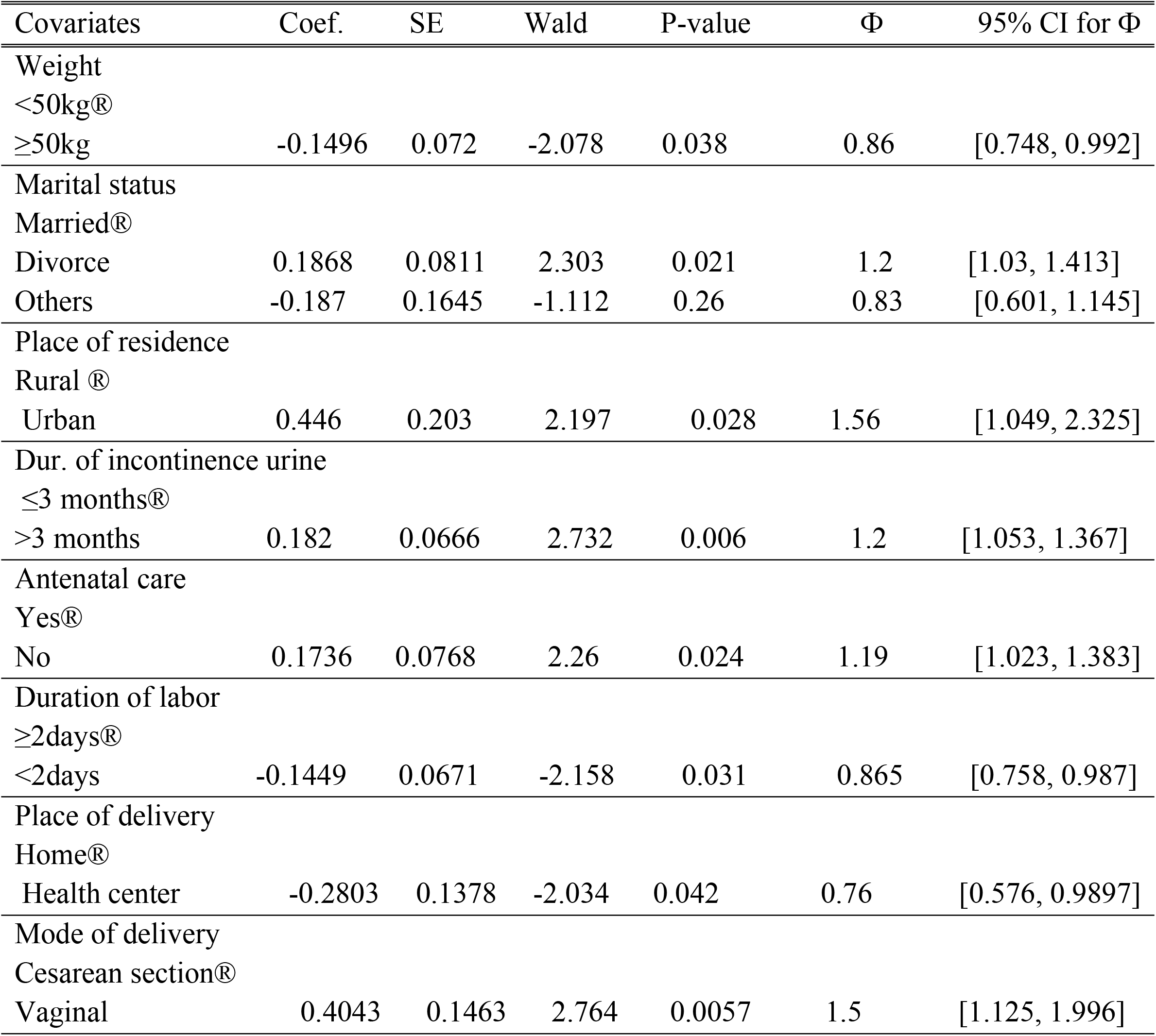
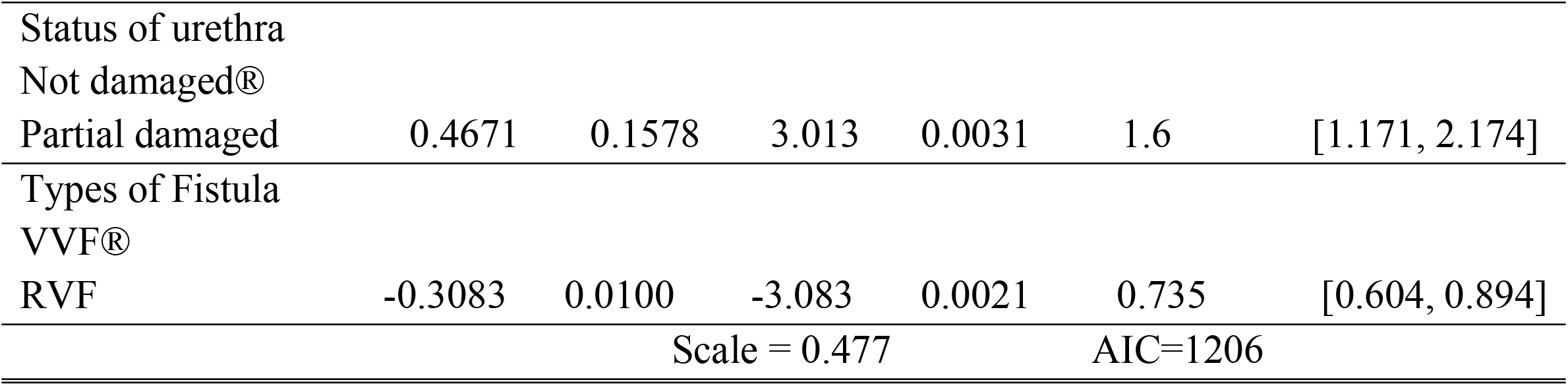
Results of multivariable lognormal AFT model for time-to-recovery from OBF

### Parametric Shared Frailty Model Results

The AIC value for both gamma and inverse Gaussian parametric shared frailty models with three baseline distributions is summarized in table 6 below. The AIC value of the lognormal inverse-Gaussian model is 1199.655, which is the minimum from all the other AIC values of the models which indicates that it is the most efficient model to describe the OBF dataset among the various parametric frailty models.

**Table 6:** AIC value of parametric frailty model for time-to-recovery from OBF in JUMC

| Baseline Distribution | Frailty Distribution | AIC |
| --- | --- | --- |
| Weibull | Gamma | 1212.534 |
|  | Inverse-Gaussian | 1209.693 |
| Lognormal | Gamma | 1204.017 |
|  | Inverse-Gaussian | 1199.655* |
| Log-logistic | Gamma | 1201.315 |
|  | Inverse-Gaussian | 1205.697 |

### Lognormal Inverse-Gaussian Frailty Model Result

Kendall’s tau (τ), which measures dependence within clusters, is estimated to be 0.074. The estimated normal is 1.527. Showed uni-modal its maximum point.

From table 7 above, showed that recovery among patients JUMC. The estimated acceleration factor for patients weight ≥50kg to less than 50kg was 0.86 with [Ф=0.86, 95% CI = 0.216, 1.656]. This suggested that the time-to-recovery for weight ≥50kg is decelerated by a factor of 0.86 compared to the weight <50kg. The estimated acceleration factor for patients’ mode of delivery at the vaginal to the cesarean section was 1.5 with [Ф= 1.5, 95% CI: 0.565, 1.645], which shows that those patient whose the recovery time for the mode of delivery with vaginal was accelerated by 1.5 times the mode of delivery with cesarean section.

**Table 7:**
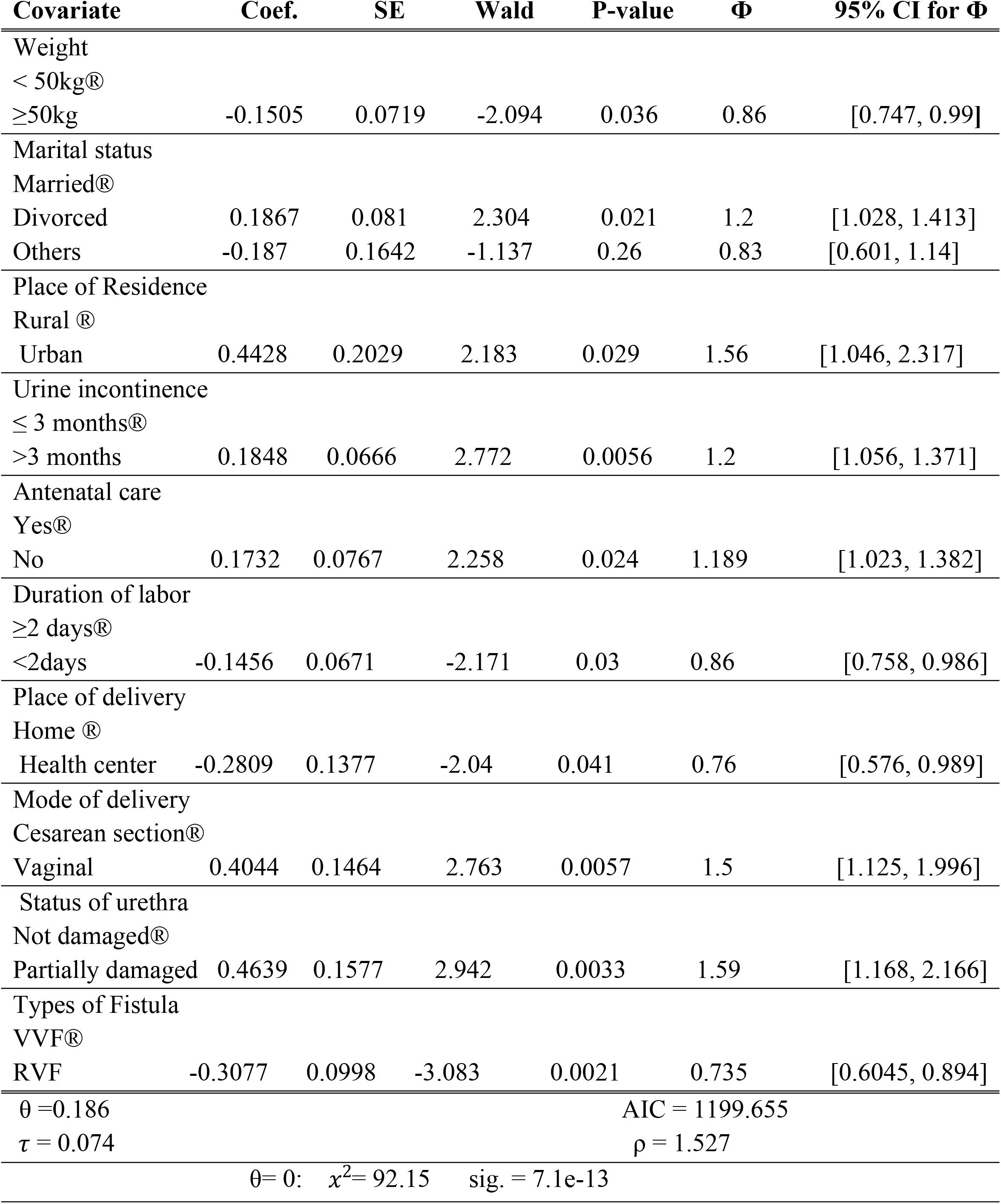
Results of Lognormal Inverse-Gaussian Frailty Model for Time-to-recovery from obstetric fistula in JUMC

## Discussion

Patients with a weight of less than 50kg has a faster recovery time than those with a weight of more than 50kg. The findings are consistent with previous research (15, 16, 17). One of the factors affecting recovery time from fistula is marital status, which suggests that divorced women have a longer time to recover from obstetric fistula than married women. A previous study backs up this conclusion (16). Living in the city accelerates recuperation by a factor of 1.56 compared to living in the country. The outcome is comparable to that of the previous study (18). Another component with accelerated factor 1.2 is the duration of incontinence of urine, which indicates that the time-to-recovery for a patient’s incontinence of urine > 3 months is longer than for a patient’s incontinence of urine 3 months. This means that the shorter the time of urine incontinence before surgical repair, the faster the recovery, and the longer the time of urine incontinence before surgical repair, the slower the recovery. This outcome is in line with (16, 17).

The recovery from obstetric fistula with prenatal treatment. Patients who did not receive antenatal care follow-up had a longer time to recover from OBF than those who did receive antenatal care follow-up. When prenatal care is used, the chances of recovery are better than if no ANC is used. Previous research backs up this conclusion (16, 17, 19). Another indicator that showed a quick recovery was the patients’ labor, which lasted just 2 days compared to the patients’ labor, which lasted 2 days. This could imply that there will be less tissue damage and a faster recovery after surgery if the obstruction is removed quickly (16, 17, 20, 21, 22). The place of birth has been found to be a major factor. When compared to delivery at a health center, the time to recovery for a home birth is longer. This outcome is similar to investigations conducted in Niger (16, 17, 23, 25).

When compared to CS delivery, vaginal birth had a longer recovery period. This can be explained by the fact that practically all OBF are caused by obstructed labor, and those who had access to CS after obstructed labor had less tissue damage from vaginal birth and could recover faster (16, 17, 23, 26, 27). Patients with a partially injured urethra take longer to heal than those who do not have a partially damaged urethra. This is due to the fact that the main continent mechanism for urine lies, resulting in surgical failure or incontinence. This confirms the findings from the earlier investigation (16, 17). The VVF has a longer survival time than the RVF, meaning that patients who have VVF have a longer recovery period than individuals who have RVF. This is because, depending on the type of vesical fistula, patients with RVF repair will be discharged home in three to five days, whereas VVF patients would be hospitalized for at least two to three weeks (28).

## Conclusion

The goal of the study was to see how long it took for obstetric fistula patients at JUMC to recover from OBF and what factors were involved. Eighty-one percent of individuals with fistulas were said to have recovered. Patients with a weight greater than or equal to 50 kg, a delivery at a health center, a labor duration of less than 2 days, and a recto-vaginal fistula had a faster recovery time from OBF, whereas urine incontinence lasting more than 3 months, urban residence, no antenatal care follow-up, vaginal delivery, divorce, and a partially damaged urethra had a longer recovery time from OBF.

## Data Availability

All relevant data are within the manuscript

## Abbreviations

AIC: Akaike’s Information Criteria
AFT: Accelerated failure time
ANC: Antenatal care, 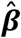: Regression coefficient
SE: standard error of 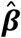
ρ: shape
σ: is the scale parameter
ε: error distribution
®: Reference group
RVF: Recto-Vaginal Fistula
VVF: Vesico-Vaginal Fistula
OBF: Obstetric fistula
JUMC: Jimma University Medical center

## Acknowledgments

The authors would like to express their gratitude to Jimma University for their financial support. We would like to express our gratitude to the Department of Obstetrics and Gynecology at Jimma University Medical Center for their unwavering support. We also like to express our gratitude to the data collectors and supervisors.

## Data Sharing Statement

The authors confirm that all the data supporting the findings of this study are available within the article.

## Ethical Approval

Ethical approval of this study was approved after review by the “Jimma University Institutional Research and Ethical Review Board,” and the research was carried out only after an ethical letter was obtained. To ensure the confidentiality of the participant’s information, codes were used instead of the names of the participant. An official letter of cooperation was taken from the respective departments and medical director of the hospital.

## References

1. Tunçalp Ö, Tripathi V, Landry E, Stanton CK, Ahmed S. Measuring the incidence and prevalence of obstetric fistula: approaches, needs, and recommendations. Bulletin of the World Health Organization. 2014; 93:60–2.

2. Tebeau PM, Fomulu JN, Khaddaj S, de Bernis L, Delvaux T, Rochat CH. Risk factors for obstetric fistula: a clinical review. International urogynecology journal. 2012 Apr 1; 23(4):387–94.

3. Adler AJ, Ronsmans C, Calvert C, Filippi V. Estimating the prevalence of obstetric fistula: a systematic review and meta-analysis. BMC pregnancy and childbirth. 2013 Dec 1; 13(1):246.

4. Hardee K, Gay J, Blanc AK. Maternal morbidity: a neglected dimension of safe motherhood in the developing world. Global public health. 2012 Jul 1; 7(6):603–17.

5. World Health Organization, UNICEF. Trends in maternal mortality: 1990 to 2013: estimates by WHO, UNICEF, UNFPA, The World Bank, and the United Nations Population Division: executive summary. World Health Organization; 2014.

6. Aalen O, Borgan O, Gjessing H. Survival and event history analysis: a process point of view. Springer Science & Business Media; 2008 Sep 16.

7. Kaplan EL, Meier P. Nonparametric estimation from incomplete observations. Journal of the American statistical association. 1958 Jun 1; 53(282):457–81.

8. Collett D. Modelling survival data in medical research. CRC Press; 2015 May 4.

9. Cox D. Regression models and lifetables (with discussions). J Roy Stat Soc 1972; 34(2):187–220

10. Schoenfeld D. Partial residuals for the proportional hazards regression model. Biometrika. 1982 Apr 1; 69(1):239–41.

11. Klein JP, Moeschberger ML. Survival analysis: techniques for censored and truncated data. Springer Science & Business Media; 2006 May 17.

12. Wienke A. Frailty models in survival analysis. CRC Press; 2010 Jul 26.

13. Hougaard P. Analysis of multivariate survival data. Springer Science & Business Media; 2012 Dec 6.

14. Akaike H. A new look at the statistical model identification. IEEE transactions on automatic control. 1974 Dec; 19(6):716–23.

15. Wall LL, Karishma JA, Kirschner C, Arrowsmith SD. The obstetric vesicovaginal fistula: characteristics of 899 patients from Jos, Nigeria. American journal of obstetrics and gynecology. 2004 Apr 1; 190(4):1011–6.

16. Feysal K. Survival Analysis 0f Time to Recovery from Vesico-Vaginal Obstetric Fistula: A Case Study at Metu Hamlin Fistula Center, Metu, South West Ethiopia; 2014

17. Getachew1 T, Ayele Taye, Jabessa, and S. Survival Analysis of Time to Recovery from Obstetric Fistula: A Case Study at Yirgalem Hamlin Fistula Hospital, Ethiopia. J Biom Biostat. 2015;6(3).

18. Muleta M, Rasmussen S, Kiserud T. Obstetric fistula in 14,928 Ethiopian women. Acta obstetrician et gynecological Scandinavica. 2010 Jul; 89(7):945–51.

19. Gessessew A, Mesfin M. Genitourinary and rectovaginal fistulae in Adigrat Zonal Hospital, Tigray, north Ethiopia. Ethiopian medical journal. 2003 Apr; 41(2):123–30.

20. Cisse CT, Faye EO, De Bernis L, Dujardin B, Diadhiou F. Cesarean section in Senegal: availability and quality of service. Cahiers d’études et de recherches francophones/Santé. 1998 Nov 24; 8(5):369–77.

21. Stekelenburg J, Schutte J, Walraven G. The use of audit to identify maternal mortality in different settings: is it just a difference between the rich and the poor. Healthcare quarterly (Toronto, Ont.). 2007; 10(2):131–7.

22. Tebeu PM, Fomulu JN, Khaddaj S, de Bernis L, Delvaux T, Rochat CH. Risk factors for obstetric fistula: a clinical review. International urogynecology journal. 2012 Apr 1; 23(4):387–94.

23. Hougaard P. Analysis of multivariate survival data. Springer Science & Business Media; 2012 Dec 6.

24. Harouna YD, Seibou A, Maikano S, Djambeidou J, Sangare A, Bilane SS. The VVF Obstetric Cause: Survey of 52 Women Admitted to the Village of Fistula. [La fistule vésco-vaginale de cause obstétricale: Enquête auprès de 52 femme’s admises au village des fistuleuses]. Médecine d’Afrique Noire. 2001; 48:55–9.

25. Sori DA, Azale AW, Gemeda DH. Characteristics and repair outcome of patients with Vesicovaginal fistula managed in Jimma University Teaching Hospital, Ethiopia. BMC urology. 2016 Dec 1; 16(1):41.

26. Stanton SL, editor. Clinical gynecologic urology. Mosby Incorporated; 1984.

27. Symmonds RE. Incontinence: vesical and urethral fistulas. Clinical Obstetrics and Gynecology. 1984 Jun 1; 27(2):499–514.

28. Tukur I, Ijaiya MA, Su TT, Chan CK, Muhammed-Baba TA, Karuthan C. Analysis of 137 obstetric fistula cases seen at three fistula centers in northwest Nigeria. East African Medical Journal. 2015; 92(8):408–14.

